# MMFP-Tableau: Enabling Precision Mitochondrial Medicine through Integration, Visualization, and Analytics of Clinical and Research Health System Electronic Data

**DOI:** 10.1101/2024.01.03.24300791

**Authors:** Ibrahim George-Sankoh, Laura E. MacMullen, Asif T. Chinwalla, Deanne Taylor, Rebecca D. Ganetzky, Katelynn Stanley, Elizabeth M. McCormick, Zarazuela Zolkipli-Cunningham, Marni J Falk

## Abstract

**Purpose:** To describe a novel data integration workflow developed to automate clinical and research electronic health system data integration and harmonization from siloed sources for centralized access, visualization and analysis by clinicians and researchers in an end user-friendly customized analytic platform.

**Methods:** A centralized, semi-automated framework provides data provenance and user access to integrated data sources. Data models are implemented leveraging a centralized server (Alteryx) for high-level analytics including scheduling, integration and modeling. A secure Tableau instance hosts end-user visualizations, with minimal software development required.

**Results:** MMFP-Tableau, named for its origin in the Mitochondrial Medicine Frontier Program (MMFP) at Children’s Hospital of Philadelphia, facilitates direct access to integrated, highly robust health system datasets. This scalable data solution enables integration of clinical and research parameters with research samples; enhances external biopharma collaborations for clinical trial design, subject recruitment and data tracking; accelerates retrospective clinical cohort data analysis; and improves complex data visualization for clinicians and researchers.

**Conclusion:** MMFP-Tableau promotes complex data integration, visibility, and advanced analytic capabilities to facilitate seamless multidisciplinary research, benefitting clinical care and research in rare disease patients and cohorts. This approach represents a generalizable workflow concept readily adaptable to implement across diverse fields of medicine

## INTRODUCTION

Data-driven precision medicine strategies are important to advance mechanistic understanding and clinical outcomes in highly individualized and heterogenous rare disorders, such as primary mitochondrial disease (PMD). Novel informatics approaches are needed to enable intuitive end-user mining of clinical data, such as from the electronic medical record (EMR), integrated with objective and subjective outcome assessments at both the individual patient and clinical cohort levels. However, data generated by health care institutions in both biomedical research and clinical domains continue to outgrow existing data management infrastructures to collect, integrate, and validate generated data from source to end-user.

Data is generated through clinical care, clinical research, and translational research to address broad operational needs. These data tend to be housed in individual siloes and in varying formats. While integrating these data is challenging due to diverse collection procedures and platforms, data compatibility and interpretability are critical requirements of accurate data mining and assessment. Clinical data from within a health system EMR are among the most complex data from a schematic viewpoint, particularly due to rich phenotypic data embedded in free text formats, as well as variable data input across different specialties and even different providers. Various processes have been utilized and evaluated to explore how algorithms incorporating EMR data can improve specific efforts such as phenotyping and defining diagnostic criteria^1^. However, the heterogeneity of different EMR database structures with respect to how datasets can be distributed and consumed remains a hurdle for accessibility and technical interoperability.

Here, we describe ‘MMFP-Tableau’, a bioinformatics platform generated in the Mitochondrial Medicine Frontier Program (MMFP) at the Children’s Hospital of Philadelphia (CHOP). MMFP-Tableau represents an innovative data warehousing framework that centrally extracts, unifies, cleans, and integrates data sources from a variety of clinical, genomic, and clinical research siloed data sources to enable end-user visualization and query within a readily customized, user-friendly platform. Integrated data sources include study demographic and clinical metrics housed in the EMR (EPIC, which is the EMR used by CHOP), research study key linking subject medical record number with study ID, and scores from standardized outcome measure electronic surveys. These data are then modeled and visualized in a secure Web environment made available to clinicians and researchers with specific permissions. Indeed, MMFP-Tableau is iteratively adaptable to requests from specific users regarding data parameters and integrated modeling to generate informative visualizations. In these ways, biomedical data integration and complex analytics modeling can help drive forward clinical care by providing supporting data to assist with making accurate care decisions, as well as research by informing longitudinal studies, preclinical evaluation of subjects’ samples, clinical trial design, precision treatment trials, and outcome measure development for PMD. The programmatic advances in clinical innovation and clinical research discovery that have been facilitated in Mitochondrial Medicine by this platform are readily translatable to other complex clinical and research populations.

This programmatic effort was initiated to provide a centralized platform to enable clinicians and researchers to directly access, interpret, and query patient-level and clinical cohort data. This, in turn, would inform patient care, facilitate queries to answer critical research questions and link human samples with accurate subject data, improve diagnostic capability, and optimize clinical trial design by better understanding cohort characteristics and enabling efficient identification of eligible patients for available clinical trials. The developed workflow integrates data from the CHOP EMR (EPIC), Research Electronic Data Capture (REDCap)^2,3^ survey software, and various Excel (Microsoft) spreadsheets located in institutional shared drives into a central server. This approach enabled a centralized data integration analyst to apply machine learning algorithms to facilitate accurate data analysis. Each independent data source contains relevant data that otherwise do not connect. For example, clinical data from the EMR is not readily viewed together with research data from interventional or observational trials. Furthermore, in multiple instances, data duplication or loss occurs due to manual curation of source data by different research personnel, given the inability to identify associations and/or download data directly from these sources. Key requirements of this integration were maintaining data integrity without duplication or loss, automating source data downloads to expedite speed and minimize personnel cost of performing individual patient chart-level manual data extraction, enabling regular streaming updates, providing selective accessibility for specific clinical and research staff, and assuring seamless connectivity between multiple sources of patient data.

## MATERIALS AND METHODS

### Description of Data Sources and Analytic Tools

Research Electronic Data Capture, REDCap, is a user-friendly data capture system that employs discrete fields for data entry. Within CHOP MMFP, REDCap is used for research data collection including recruitment and enrollment tracking, research cohort delineation, subject genetic etiologies, observational research data (objective and patient-reported data), and laboratory specimen data (including locations and descriptions of biological samples).

EPIC, the EMR system currently utilized by CHOP, houses all clinical data pertaining to patient care including, but not limited to, demographics, physical exam descriptions, laboratory test results, medications, phenotypes and genotypes, specialty assessment information, hospitalization and infection history, and imaging. All data in EPIC is directly accessible through a back end data warehouse, Clarity.

Data extraction, cleaning, and visualization were completed using two customizable off-the-shelf data platforms, Alteryx Version 2021.4.2.47844 and Tableau Version 20222.23.

*Alteryx* is an analytics platform that enables creation of analytical workflows with minimal programming and development of reusable macros and templates. The Alteryx Scheduler Tool enables scalable computational resources for processing large datasets and scheduling real-time updates from live data sources. The advantage of this tool is to provide a plug-and-play interface that reduces the need for a full team of informaticians and data engineers, allowing a data analyst to easily build and customize data workflows. Using Alteryx, rapid iterations of Quality Improvement (QI) can be performed.

*Tableau* is a commercial software platform for developing interactive dashboards for data visualization, supporting approved data users to query, view, and download data subsets for further analysis. The secure role-based access functionality enables controlled user access to identifiable patient data protected by the Health Insurance Portability and Accountability Act of 1996 (HIPAA), and specific research protocols. The CHOP Tableau instance is centrally administered behind a secure institutional firewall.

### Data Extraction

Extraction of clinical data was limited to patients evaluated in the CHOP MMFP. Any data used for subsequent research analyses was further filtered to include only patients consented to an active CHOP IRB-approved study protocol (#08-6177, Falk PI). EMR clinical data extracted from Clarity included both structured and narrative data, clinical flowsheets, and scanned PDFs. Clinical data is entered into EPIC in the context of clinical care by mitochondrial medicine and specialist physicians as well as other ancillary clinicians including genetic counselors, physical therapists, nurses, and dieticians. Clinical data was pulled from Clarity tables using Structured Query Language (SQL). Macros within Alteryx were developed and run on the textural fields to extract specific words and discretize narrative data. Phenotype data was mapped with human phenotype ontology (HPO) and ICD10/SNOMED codes to facilitate extracting discrete elements that describe coded diagnoses.

Research data was originally captured from a mixture of excel spreadsheets and REDCap. To reduce issues with data formatting, consistency and version control, research data collection was programmatically standardized to use structured templates in REDCap. These REDCap databases, primarily used for collection and organization of research and laboratory data, were pulled into Alteryx using an Application Programming Interface (API). REDCap is role-based for both data entry and API access^3^. Alteryx has a scheduler to automate data extraction and analysis workflows, thereby eliminating the need for manual extraction and providing real-time data from clinical and research data sources. Extracted data is stored and visualized in Tableau for end-user analysis and feedback.

### Data Quality

An interdisciplinary team of clinicians, researchers, analysts, and scientists reviewed data visualizations on an ongoing basis to identify data discrepancies, prioritize data selection, and develop strategies to optimize data visualization for maximum utility. This iterative process also supported programmatic restructuring of data input by allowing clinicians and researchers to see the value of creating and using discrete data fields in clinical care documentation entered into EPIC. For example, while phenotype data was often outlined in textural clinical notes, the data extraction process made clear that careful use of diagnostic codes within the structured ‘problem list’ in EPIC allowed for structured data to be most reliably used for accurate cohort-level analyses.

### Data Transformations and Analysis Methods

Our Extract, Transform, and Load (ETL) process, shown in **Figure 1**, involves extracting data from siloed sources and transforming it to fit programmatic needs. The staging platform is used to join data sources, assign variable names, and apply database constraint rules to eliminate duplicate entries. This high-level structure enables agile development of data transformation workflows for data extraction and loading large volumes of data inputted from multiple sources. The analytics team worked closely with MMFP clinicians to define inclusion and exclusion criteria for patient phenotype categories based on clinical features. The Alteryx platform includes tools for data preparation, integration, cleaning, modeling, and predictive analytics. Inbuilt statistical regression modules were implemented for analytical workflows, including machine learning methods.

**FIGURE 1:**
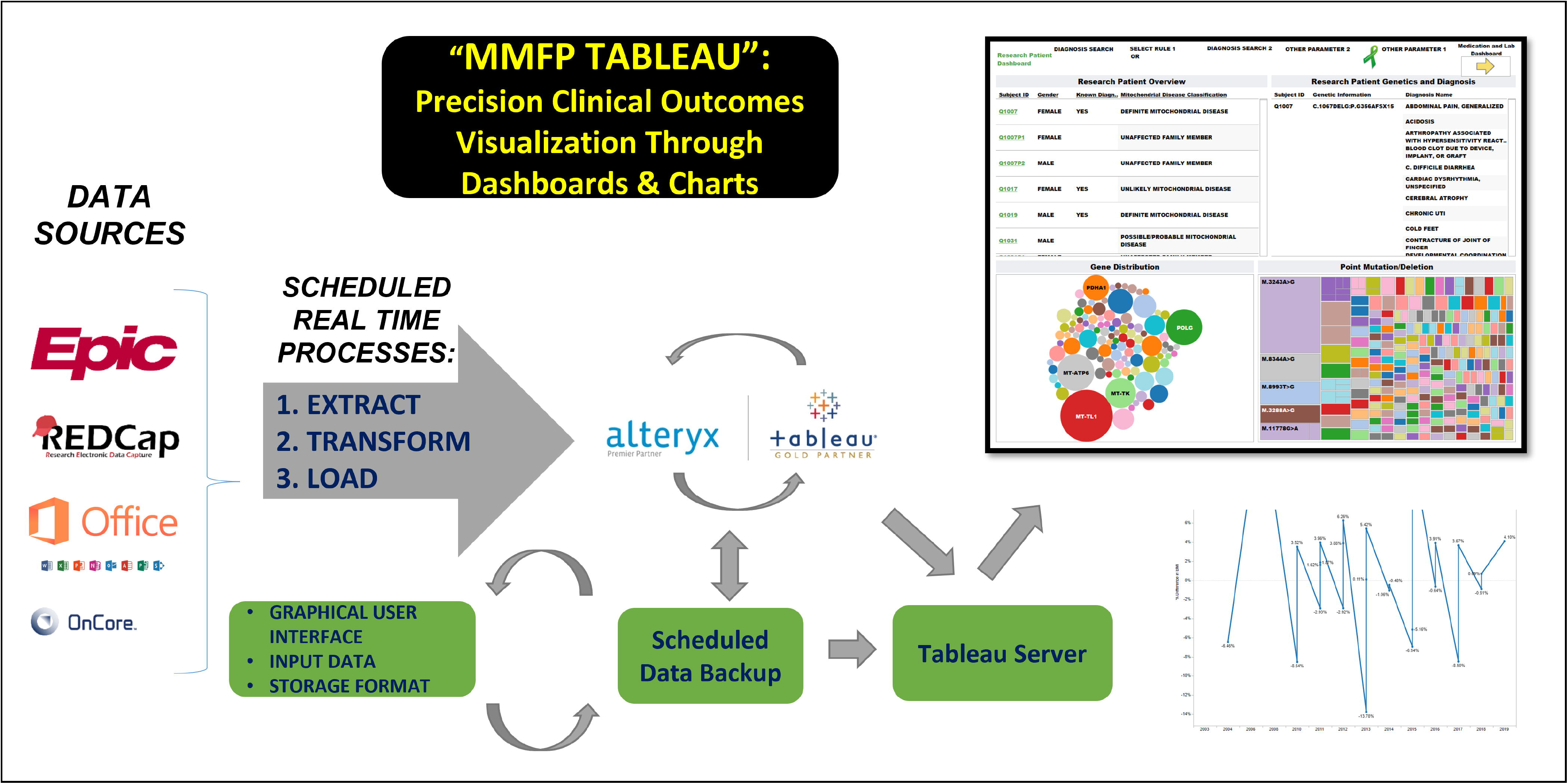
Schematic Overview of Mitochondrial Medicine Frontier Program (MMFP) Data Integration Platform Methodology. The MMFP-Tableau data platform integrates multiple data sources using real-time, scheduled Extract, Transform, Load (ETLs) processes that are processed first through the secure Alteryx server and finally into Tableau for interactive, dynamic, visualizations through dashboards and charts, and advanced queries by approved clinician and research end-users.

### Data Visualizations

Analyzed data and visualizations were presented to pilot users throughout the development process to improve MMFP-Tableau user interfaces and cross-check the accuracy of extracted data. Incremental rollout of the MMFP-Tableau system allowed for ongoing system improvement. The customizable visualization tools of the MMFP-Tableau platform support access to controlled data dashboards, dynamic filters, custom queries, and statistical regression analytic methods by end-users.

## RESULTS

MMFP-tableau integrates data from over 2,000 individuals evaluated in the CHOP MMFP clinical program and/or enrolled in research, including EMR (clinical notes and/or discrete problem lists, clinical laboratory results, etc.) and REDCap (patient-reported outcomes, observational research, human samples) data. The model uses structured tables processed from raw data extracts to enable performance by clinicians and researchers of complex data visualizations and analyses within the MMFP-Tableau custom environment (**Figure 1**). Staging processes are structured in layers to ensure maintainability, reusability, provenance, and scalability. Workflows are optimized at every level of integration by implementing parallel processing, where all extractions are performed individually on specific data modules.

Queries are enabled by live filters and the aggregated macros from the staging platform that can return different view counts and percentages. These views can drill down to granular points for end-users exploring the data. Depending on access levels granted, MMFP-Tableau users can also adapt filters and views to meet their current needs (**Figure 2**), including: (a) a cohort-level view with full patient list (accessible by medical record number or study ID), ICD-10 diagnoses, distribution graphs of causal gene(s) and pathogenic variants (point mutations or deletions), and free text search filters to allow streamlined queries of patient subgroups; (b) individual patient laboratory test results and medications; and (c) individual patient data including hospitalizations and patient-reported outcome (PRO) electronic REDCap survey data; (d) locations and volumes of research study subject samples stored in the MMFP Research Laboratory. Each view pulls live data in real-time as continuous updates are made to sources.

**Figure 2:**
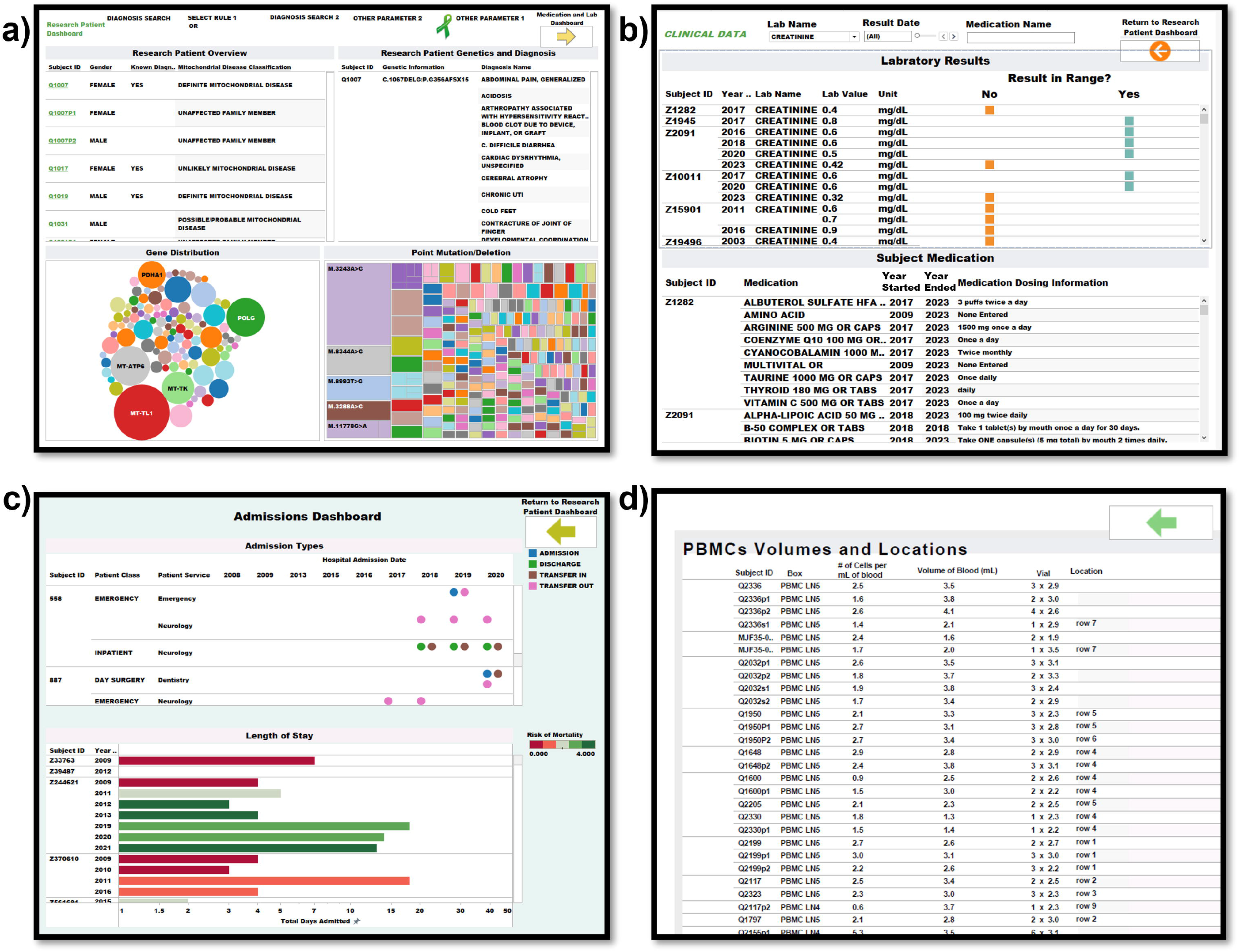
MMFP-Tableau Clinical Research Dashboards. MMFP-Tableau user interface examples are shown that include linked dashboards of (a) cohort-level data including diagnosis status, demographics, problem list, and pathogenic gene and mutations, (b) subject-level data including lab results and current medications, (c) subject-level data including hospital admissions, length of stay, and mortality status, and (d) locations and volumes of research study subject samples, such as peripheral blood mononuclear cells (PBMCs), stored in the MMFP Research Laboratory.

### Use cases

The following MMFP-Tableau use cases demonstrate how this novel health data visualization and analysis platform can be used to query complex and siloed health system data often entered into the EMR on a per patient basis to answer complex clinical and research questions that require integrated data parameter analyses, at both the individual patient and broader disease cohort levels. This capability enables more rapid, automated, and accurate recognition of data patterns among patients and subgroups, and directly supports a wide range of cohort-level data mining applications for purposes of research queries and clinical research subject recruitment. This system further supports standardized assessment of key clinical health system performance metrics for a target population, such as demographic trends (zip code analysis, age trends, etc.), test utilization volumes, etc.

1. *Clinical cohort characterization.* A more complete exploration of the MMFP-Tableau resource specifically for visualization and expedited analysis of PRO research survey data has been recently described ^4^. A user example for this interface is a physician-scientist seeking to characterize the prevalence of seizures in Leigh syndrome spectrum ^5^, the most common pediatric PMD clinical syndrome. In this scenario, Dashboard 1 would be searched for “Leigh syndrome” and “seizures”, which would yield the full list of MMFP Leigh syndrome spectrum patients having any clinical manifestation of seizures. The list can be further filtered by age or genetic etiology, which can be further interrogated to evaluate medications for current treatment of seizures and recent bloodwork results at the individual subject level. Finally, the user can obtain an even deeper view of the patient by reviewing recent hospital and ED admissions and PRO scores ^4^ across clinical domains of quality of life (measured using the Pediatric Quality of Life Inventory Generic Core Scales ^6,7^), fatigue (measured using the Modified Fatigue Impact Scale ^8^), and function (measured using the Lansky Scale ^9^ and the Karnofsky Scale ^10^).
2. *Clinical trial eligibility determination*. Another user example for MMFP-Tableau is ascertaining patients meeting inclusion criteria for clinical drug trials (**Figure 3**, PMD cohort subset dashboard displaying demographics and phenotype categories). This view provides an example of how consolidated cohort data with customizable filters can be used to readily identify eligible patients for available clinical trials. Users can customize the filters to the specific study inclusion and exclusion criteria, including age and other demographic factors, ambulatory status, and/or survival status. It would also be possible to filter out exclusionary criteria (not shown), such as by laboratory values or systemic features. This figure expands on clinical demographic data by also displaying ambulatory status, defined as the ability to take 5 independent steps, which represents a crucial datapoint needed to determine eligibility for recruitment into clinical trials involving myopathy or exercise capacity assessments but which is often difficult to ascertain directly from clinical EMR documentation.
3. *Genotype-phenotype mapping.* In this example, a user can visualize the distribution of signs and symptoms across the patient cohort, as categorized by causal disease gene in each patient (**Figure 4**). This type of view is helpful to characterize patient phenotypes and recognize common symptoms among each PMD cohort subtype. This view incorporates mapping of ICD-10 codes from the EMR problem list to common HPO terms that facilitate data harmonization and cross-institutional collaboration.
4. *Medication trend visualization*. In this example, a user can query the most common medications prescribed to PMD patients as categorized by therapeutic class and patient phenotype (**Figure 5**). This visualization enables systematic evaluation of prescribing patterns by different physicians and across clinical syndromes. In addition, this visualization facilitates queries of concomitant or exclusionary medications for clinical trials. Models like this can be readily adapted across multiple use cases. For example, once medication class categories are initially defined and mapped to the appropriate medications, the workflow can be modified and adapted for different uses, such as to look at medication classes by symptom or age category.
5. *Risk factor delineation*. In this last example, users can rapidly synthesize clinical cohort data to categorize specific risk factors relevant for PMD patients, such as a) risk for kidney failure as indicated by Glomerular Filtration Rate (GFR) and creatine levels, and b) biochemical laboratory test results, such as lactate, in a research sub-cohort ().

**Figure 3:**
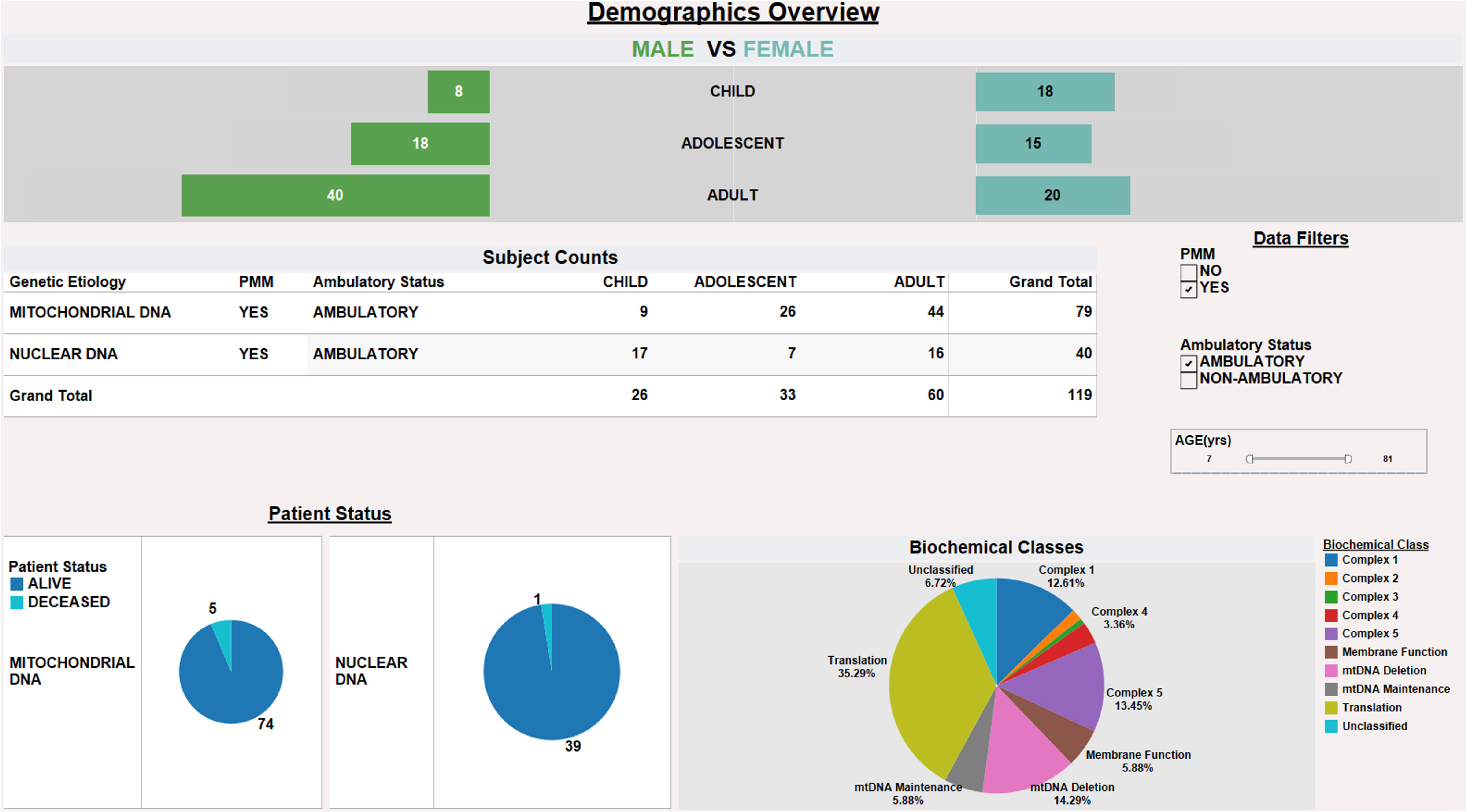
Enhanced Data Visualizations to Optimize Clinical Trial Recruitment. Dashboard view of cohort sub-analyses is shown that combines data from clinical and research sources, including demographic, survival status, and biochemical class categorization of PMD. Dynamic filters allow users to readily discern potentially eligible clinical trial participants for a given study based on study-specific inclusion and exclusion criteria including presence of Primary Mitochondrial Myopathy (PMM) diagnosis, age boundaries, and ambulatory status.

**Figure 4:**
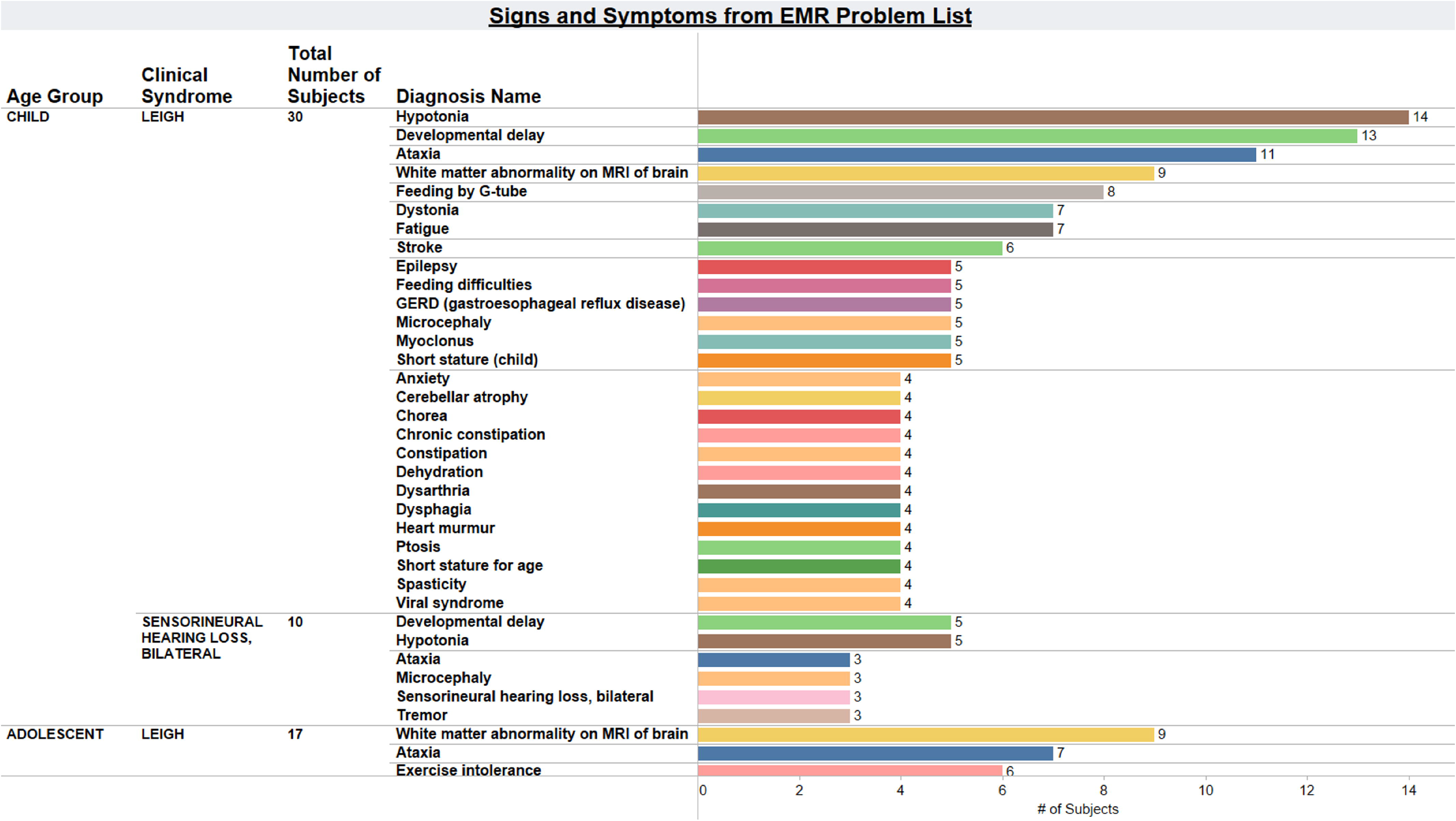
Signs and Symptoms Categorized by Clinical Syndrome. Visualization generated in MMFP-Tableau is shown that represents the prevalence of each symptom, as listed in the problem list of the EMR (EPIC) for the Leigh syndrome spectrum PMD cohort. Each symptom is an HPO term mapped to an underlying ICD10code.

**Figure 5:**
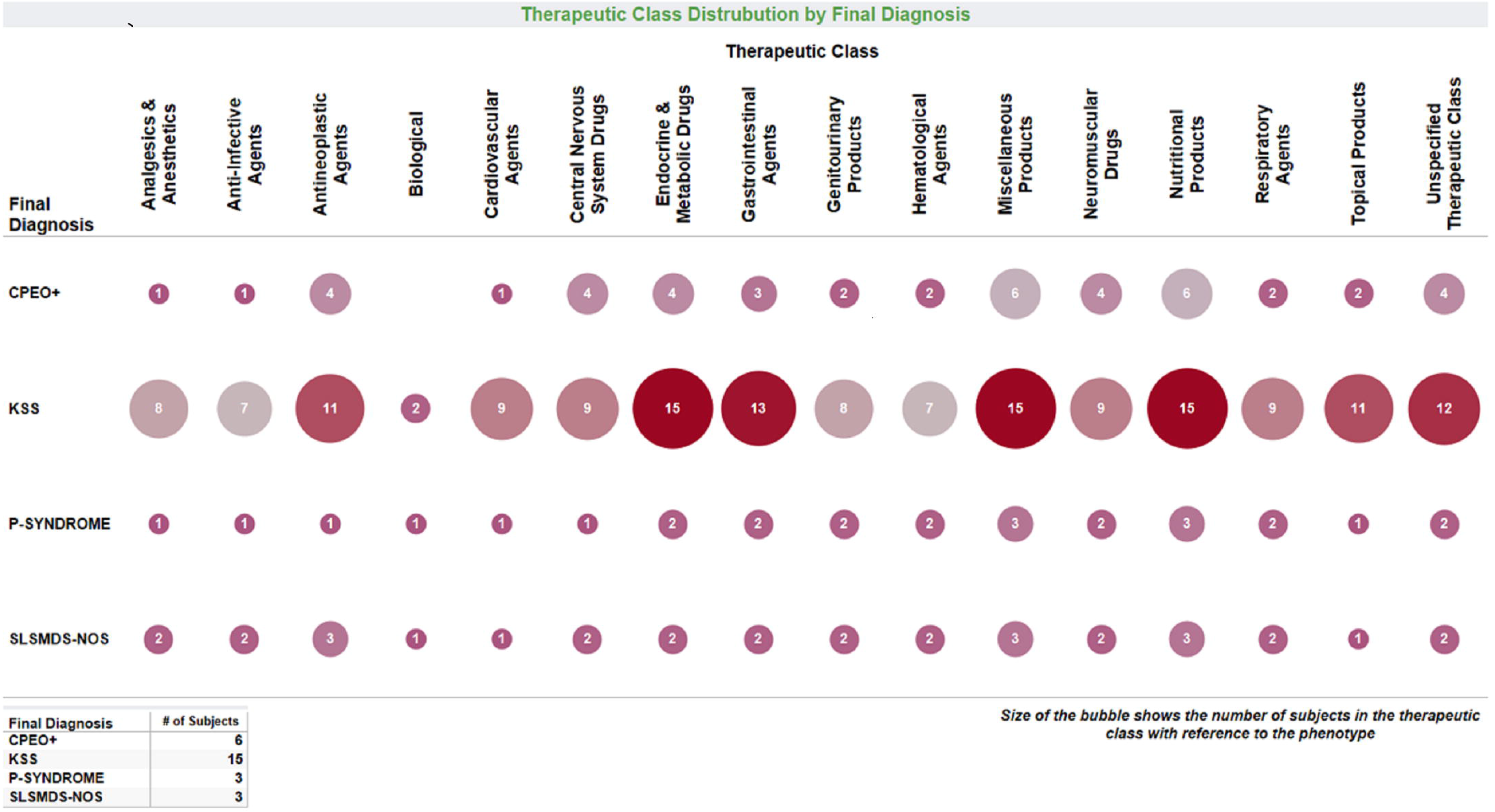
Therapeutic Classes for PMD Research Study Subject Medication Usage. Size and color show the number of subjects currently taking medications within each therapeutic class, as displayed for each PMD clinical syndrome. CPEO, chronic progressive external ophthalmoplegia. KSS, Kearns Sayre Syndrome. P-syndrome, Pearson syndrome. SLSMD, single large-scale mtDNA deletion syndrome. NOS, not otherwise specified.

## DISCUSSION

Here, we have described a novel health data integration platform, MMFP-Tableau, that extracts and synchronizes a wide array of clinical and research data elements to provide clinically meaningful and potentially actionable data for direct use by clinicians and researchers. The MMFP-Tableau platform places real-time, synthesized, accurate data pulled directly from the source at the fingertips of clinician and researcher end-users, bypassing the need for sophisticated informatics and data modeling expertise while streamlining infrastructure support, enabling novel clinical and research insights to be made and rapid interrogation of longitudinal health system data at both individual patient and cohort-wide levels.

The MMFP-Tableau platform provides a new paradigm to address the challenges of reviewing large-scale electronic health system data in a cost-effective fashion. Indeed, it has transformed manual *ad hoc* processes such as serial manual chart reviews into a highly efficient, systemized, and reproducible framework. Harnessing this informatics capability enables the identification of potential subjects for clinical trial enrollment, provides accurate data to inform improved biopharma-driven patient registries and clinical trial design, enables research biospecimen tracking in an integrated fashion with relevant phenotypic information, standardizes phenotyping in a defined HPO ontology among medically complex patients, and allows sophisticated characterization of diverse aspects of clinical presentations in a genetically and phenotypically heterogeneous rare disease. Perhaps most importantly, MMFP-Tableau has unlocked a reservoir of available clinical and research healthy system data that can initiate and support new collaborations between academic institutions and pharmaceutical companies to optimize the therapeutic development process and clinical trial design based on timely access to real-world data.

Studies have found that on average physicians spend 15-38% of their time in direct patient care and 50-67% of their time on indirect care ^11–15^, including review of electronic medical records (EMRs) documentation and navigation of multiple data sources to input or review new results. Having data readily accessible for clinician queries in pre-determined dashboards within MMFP-Tableau increases both time and cost efficiencies, and may also help inform clinical care decision-making capacity, by providing clinicians with more accessible individual and cohort-level data in an integrated and end user-friendly interactive analysis format.

Importantly, clinician use of the MMFP-Tableau system has reshaped EMR clinical documentation and data literacy of team members within the MMFP. As outputted data became more readily available for extraction, analysis, and visualization, clinicians were able to recognize first-hand the utility of having discrete data elements included in their clinical documentation, which could be readily extracted to enhance discovery of complex patterns across patient cohorts that may not be readily apparent when thinking about patients on a case-by-case basis in clinic. Indeed, within our clinical group, EMR templates have been systematically improved to include more discrete data fields that capture causal gene etiologies, specific pathogenic variants, and mitochondrial DNA variant heteroplasmy levels.

The MMFP-Tableau Web platform interface is readily accessible for end-user analysis of both identifiable (linked to medical record number) and de-identified (linked to research study ID) data, depending on approved user roles. It streamlines data capture and analytic workflows, while increasing accuracy and integrity of the generated datasets. With patient consent, this data platform unifies EMR data (including genetic information, complex phenotypic data from problem lists and physical exam findings, medications, laboratory test results, imaging findings, and ancillary assessments such as physical therapy quantitative metrics), curated research data (including observational, interventional, and translational research data), and a potentially unlimited array of other siloed health system clinical and research data sources that do not typically communicate. Integrated data is harmonized and presented within MMFP-Tableau in a custom dashboard to support end-users’ ability to make dynamic insights into patient and cohort-level clinical data.

Harmonizing language for entry of data inputted across medical centers, or even within the same medical center, remains a challenge, particularly when describing patient phenotypes. The use of Human Phenotype Oncology (HPO) codes has proven vital to enable collaborative research in medicine ^16^. When conducting multi-center studies, particularly international studies, it is imperative that clinicians and researchers utilize the same language to discuss patient phenotypes. From an analytics perspective, HPO codes allow for discretized data and 1:1 mapping when comparing or modeling phenotypes for diagnostics. For these reasons, the MMFP-Tableau platform adopted HPO codes to facilitate future data interoperability between centers.

Data integrity has profound consequences in terms of quality and use. As even small-scale health system projects may generate massive usable data, challenges remain in stewardship or ownership to maintain data integrity. For instance, limited harmonization exists of data structure across research datasets, which often depend exclusively on the medium of data collection used. Even when utilizing metadata for summarizing data content within a source, wide variability exists in available source documentation necessary for accurate relationship mapping. Maintaining data provenance and data integrity is crucial to data credibility when analyzing large, disparate datasets. Data management and integration platforms have not yet yielded consistent solutions for these challenges, where new questions continually arise about database design structure, data extraction, and proper query structuring. It is in this context that MMFP-Tableau provides an innovative and transferable solution to extracting, harmonizing, relationship mapping, and integrating complex health system data in a systematic, structured fashion that is clinically-meaningful and readily accessible to end-users.

Accessibility to health system data in a secured and consumable format for data feedback from subject matter experts enables their access to the highest level of data integrity and quality. Due to the sensitivity of clinical and research data, considerations such as patient privacy and research regulations regarding data access, secured, and well-designed levels of access are essential. MMFP-Tableau addresses these challenges, by creating a common interface accessible by all approved clinician and research end-users that is located behind a secure institutional firewall. Further, the underlying data warehouse schema enables complex data queries and dynamic visualization of integrated data parameters while creating a level of separation between the user and the raw data source.

Current limitations of the MMFP-Tableau platform include inaccessibility of some EMR data including non-discretized data, some imaging data that are housed separately (e.g., picture archiving and communication system (PACS)), and scanned laboratory results (i.e., PDFs not able to be parsed). Our system is also currently unable to automate the extraction of data pulled into the EMR by external institutions; this remains an important future development goal to support integrated data visualizations of Fast Healthcare Interoperability Resources (FHIR) endpoints both within MMFP-Tableau and to allow broader implementation of MMFP-Tableau at outside institutions. Furthermore, the volume of live data running through the MMFP systems can cause slow server operations and recurrent technical difficulties; strategies are under development to increase system efficiencies and streamline computational processing. Ongoing work is focused on developing models for predictive diagnostics to assist in the identification of patterns in phenotypes, natural history, and potential disease biomarkers of clinical relevance to the therapeutic drug development process. The MMFP-Tableau data model can be further scaled for cohort expansion and new data fields and data types based on the platform’s streamlined and generalizable framework.

## CONCLUSION

MMFP-Tableau is a novel health data integration, visualization and integration platform developed to support programmatic implementation of precision medicine in clinical care, and seamless integration with clinical research, to enable ongoing clinical care optimization and support therapeutic development in heterogeneous rare diseases such as PMD. A centralized server (Alteryx) managed by a dedicated data integration analyst efficiently blends and models data from multiple different clinical and research sources using high-powered predictive, spatial, and statistical analytics. Modeled data are then pushed to a local instance of a commercial data visualization interface (Tableau) to support dynamic, interactive data visualization and analyses by approved researchers and clinicians. This platform supports iterative improvements in usability, with limitless potential to incorporate new data types, create new data views, and extend to new clinical cohorts. The MMFP-Tableau data integration platform has created direct end-user access to previous siloed data that are now centralized in a regularly updated fashion within a shared Web interface that connects and standardizes complex clinical and research data types. Overall, the MMFP-Tableau platform enables adaptive data queries and customizable, dynamic views using a scalable and generalizable paradigm that was built within a single health system for a single clinical and research group, but can be readily replicated to support a continual learning health system approach to improve care and research capabilities for any disease type or cohort of interest.

## Data Availability

Data and code files are available from corresponding author upon request.

## Acknowledgments

We are grateful to all the patients and families for their participation in CHOP Mitochondrial Medicine Frontier Program (MMFP) clinical care and research. We would like to acknowledge Batsal Devkota, PhD, for his work on the early pilot data extractions and workflow configurations that helped to provide the foundation for this project.

## Contributors

Conceptualization: M.J.F.; Database Creation: I.G.S.; Funding Acquisition: M.J.F.; Source Data Curation and Data Visualization Optimization: L.M., E.M.M., K.S., R.G., Z.Z.C; Methodology: I.G.S., A.C., D.T., L.M.; Writing-original draft: I.G.S., L.M., A.C., D.T., M.J.F. Manuscript Figures: K.S. All authors reviewed and approved the final manuscript.

## Ethics Declaration

All human subjects research was performed under a Children’s Hospital of Philadelphia (CHOP) Institutional Review Board approved study #08-6177 (Falk, PI).

## SUPPLEMENTARY FILE LEGEND

**Supplemental Figure 1:**
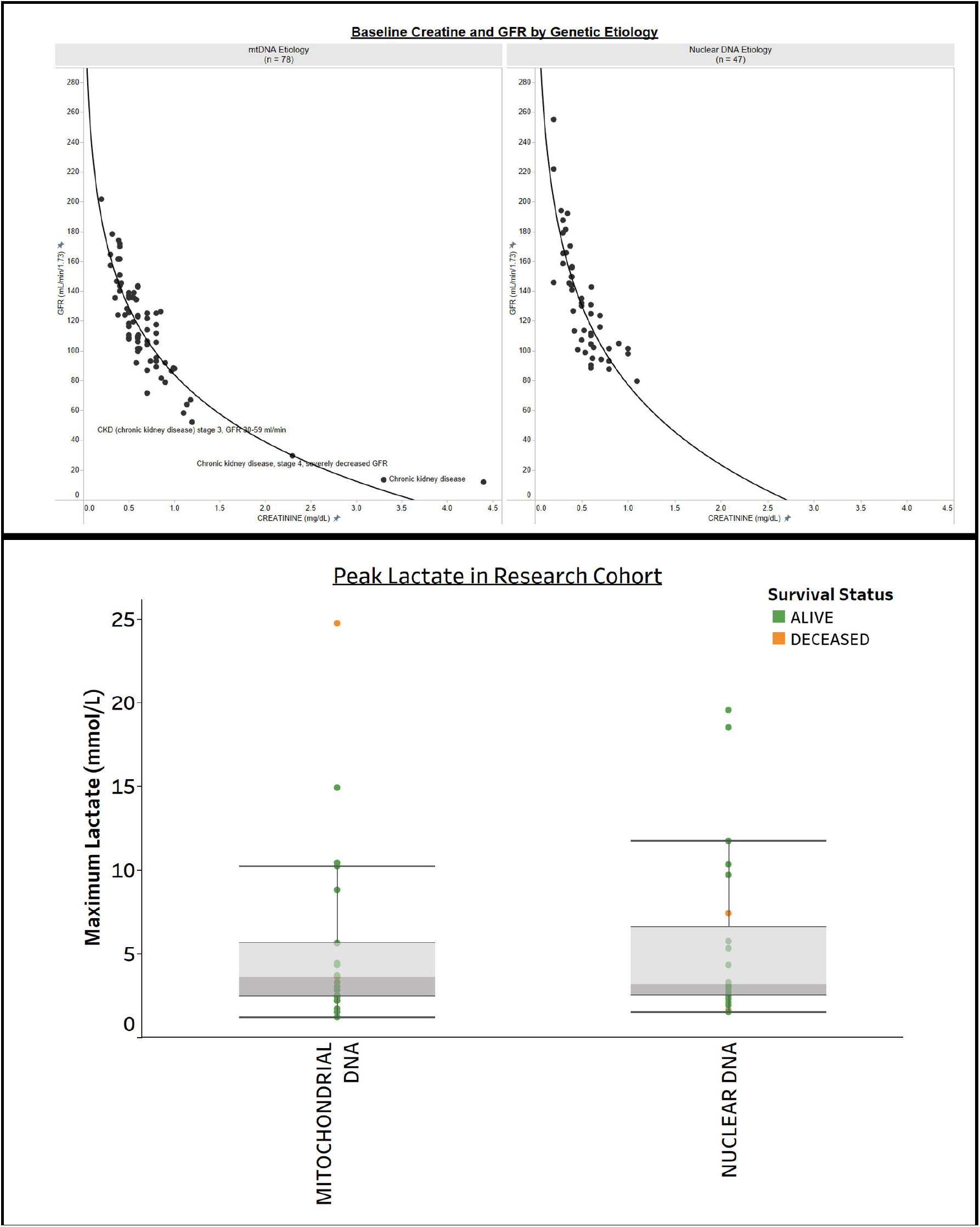
Visualization of Laboratory Biomarkers in PMD subcohorts. **(a) <u>Creatinine and glomerular filtration rate (GFR) by Genetic Etiology.</u>** Glomerular Filtration Rate (GFR) shows renal filtration capacity. This view allows clinicians to visualize and explore risk factors for kidney failure in PMD for specific mitochondrial DNA (mtDNA) or nuclear DNA genetic etiologies, and perform analyses of statistical correlations. **b) <u>Biochemical Laboratory Test Results: Lactate.</u>** The maximum lactate level for each subject was pulled from the medical record in a PMD sub-cohort of 47 subjects aged 0 to 31 years. This display further categorizes subjects by their mitochondrial or nuclear DNA gene etiology and survival status (green and orange, respectively, indicates the subject was alive or deceased at the time of the source data pull into MMFP-Tableau).

